# The Association between Multimorbidity and Out-of-Pocket Expenditures for Prescription Medicines among Adults in Denmark: A Population-Based Register Study

**DOI:** 10.1101/2023.11.21.23298458

**Authors:** James Larkin, Susan M. Smith, Line Due Christensen, Thomas Schmidt Voss, Claus Høstrup Vestergaard, Amanda Paust, Anders Prior

## Abstract

**Background:** Multimorbidity, defined as two or more chronic conditions in an individual, is increasing in prevalence and is associated with polypharmacy. Polypharmacy can lead to increased out-of-pocket payments for prescription medicines. This, in turn, can be associated with cost-related non-adherence and impoverishment. Healthcare in Denmark is mostly free at the point-of-use; prescription medicines are one of the only exceptions.

**Objective:** To examine the association between multimorbidity and annual out-of-pocket prescription medicine expenditure for adults in Denmark.

**Methods:** A population-based register study was conducted. The study population included all adults residing in Denmark in 2020. Frequencies and descriptive statistics were used and regression analyses were conducted to assess the association between multimorbidity and annual out-of-pocket prescription medicine expenditure, while controlling for demographic and socioeconomic covariates.

**Results:** Overall, 1,212,033 (24.2%) individuals had multimorbidity. Individuals with five or more conditions spent, on average, €320 in out-of-pocket prescription medicines expenditure compared to €187 for those with two conditions and €44 for those with no conditions. Amongst those with any out-of-pocket prescription medicine expenditure, having multimorbidity was associated with 2-4 times greater out-of-pocket prescription medicine expenditure than those with zero conditions. Amongst those in the quantile with the highest expenditure, those with five or more conditions spent €408 more than those with no conditions, and those with two conditions spent €185 more than those with no conditions.

**Conclusions:** For adults in Denmark, multimorbidity was associated with significantly higher out-of-pocket prescription medicine expenditure, even after controlling for demographic and socioeconomic covariates. This is similar to patterns in other countries and likely affects those with lowest income the most, given the known socioeconomic patterning of multimorbidity and raises concerns about cost related non-adherence. Potential protective mechanisms could include subsidies for certain vulnerable patient groups (e.g. those with severe mental illness) and low-income groups.

## 1. Introduction

In most member countries of the OECD, including Denmark, there has been an increase in per capita pharmaceutical expenditure between 2011 and 2021.^1^ These trends are predicted to continue into the future.^2^ The reasons behind these increases in expenditure include the ageing of populations,^3^ rising chronic disease prevalence,^4^ an increase in prescription medicines use^5^ and the development of expensive novel medicines.^6^ Another major driver of prescription medicine expenditure is polypharmacy; the concurrent use of five or more medications by an individual. In Denmark, the prevalence of polypharmacy is considered high: 33% among those aged 65 years or over,^7^ rising to over 50% among those aged 85 years or over.^7^ In a study of 17 European countries (including Denmark), polypharmacy prevalence overall is estimated at 32% among those aged 65 years or over.^7^ Polypharmacy is relatively common in people with multimorbidity (diagnosis of two or more chronic diseases in an individual).^8–10^ Multimorbidity prevalence in Denmark in the population aged 50 years and over is estimated at 42% using self-report data from 2015I.^11^ To put this in context, a study involving Denmark and 12 other European countries, using the same 2015 self-report data, estimated the overall prevalence of multimorbidity amongst those aged 50 years and over at 48%.^11^

People often have to pay some form of out-of-pocket fee to access prescription medicines.^12^ In many countries, out-of-pocket expenditure on prescription medicine is the largest category of out-of-pocket healthcare expenditure for people with multimorbidity.^13–15^ A systematic review of 14 studies from North America, Australia and Asia, found that multimorbidity is associated with five to ten times higher out-of-pocket medication payments than for those with one chronic condition or no chronic conditions.^12^ This is primarily driven by the polypharmacy associated with multimorbidity.^12^

High out-of-pocket prescription medicine expenditure is problematic for a number of reasons. Firstly, out-of-pocket medication costs, even small costs, can lead to cost-related non-adherence.^12,16^ Secondly, out-of-pocket healthcare costs can lead to impoverishment.^12^ And thirdly, these issues are particularly problematic for people with multimorbidity, not only because they experience higher out-of-pocket costs, but also because they are more likely to have lower incomes and experience socioeconomic deprivation, with those in the most disadvantaged communities developing multimorbidity at a younger age and also experiencing more complex mixtures of physical and mental health conditions.^17–19^ The lower average income among people with multimorbidity compounds their high out-of-pocket healthcare expenditure by increasing financial burden^13,14^ – the percentage of household income spent on healthcare.

### 1.1 Health coverage in Denmark

Healthcare in Denmark is mostly free at the point-of-use.^20^ Prescription medicines are one of the only healthcare costs that are not covered by the Danish state though safety nets and upper limits to expenditure apply.^20,21^ Full details of relative entitlements in the Danish healthcare system are available in Section 2.2.1. Prescription medicines are estimated to account for one third of out-of-pocket healthcare costs.^22^ While there are built-in protections to address prescription medicines costs, out-of-pocket prescription medicine expenditure is estimated to take up 43% of out-of-pocket healthcare expenditure for those in Denmark aged 50 years or over.^11^

### 1.2 Aim

Given the increasing expenditure on medicines, the high rates of polypharmacy in Denmark, and the potential for negative effects arising from out-of-pocket prescription medicine payments, we aimed to 1) describe out-of-pocket prescription medicine expenditure and its associated financial burden for people with multimorbidity and 2) examine the association between multimorbidity and annual out-of-pocket prescription medicine expenditure for adults in Denmark.

## 2. Methods

A population-based register study was conducted. The study population for this analysis includes all adults resident in Denmark for any period between January 1^st^ 2020 to December 31^st^ 2020. This includes those based in residential care facilities.^23^ The study is reported according to the STROBE statement.^24^

### 2.1 Data

We used data from four Danish nationwide registries: *1) The Danish National Prescription Registry*,^23,25^ 2) *The Danish National Patient Registry*,^26^ 3) *Statistics Denmark*^27^ and 4) *The Danish Civil Registration System*.^28^ Matching across databases was facilitated by an individual’s civil registration number which is unique, permanent and assigned to every individual.^28^ The Danish National Prescription Registry contains information for all prescriptions redeemed in community pharmacies in Denmark since 1995.^23,25^ The Danish National Patient Registry^26^ contains information on chronic condition diagnoses in secondary care. Diagnoses made in primary care are not available from national registries. Statistics Denmark contains information on education level, population density, immigration status and household income. ^27,29^ The Danish Civil Registration System contains information on age, cohabitation status and gender.

#### 2.1.1 Multimorbidity

Multimorbidity is defined as the coexistence of two or more chronic conditions in an individual. In Denmark, diagnoses made in primary care are not available from national registries. Instead, the Danish Multimorbidity Index Algorithm^30^ (appendix A, eTable 1) was used to determine the number of chronic conditions each individual had. This uses secondary care diagnoses (using ICD-10 codes) and redeemed prescription details (using ATC codes) to determine if participants have any of the 39 conditions included in the list outlined in Appendix A. Information on chronic condition diagnosis was gathered on 1^st^ January 2020.^26^

#### 2.2.1 Out-of-pocket prescription medicine expenditure and financial burden

The primary outcome variable was annual out-of-pocket prescription medicine payments, which consists of the total out-of-pocket payments for prescription medicines made in 2020 by adults in Denmark, accounting for relevant government subsidies. Details of the calculation process for those residing in Denmark for less than 12 months or having died in the given year is in Appendix A, eBox 1.

The out-of-pocket prescription medicine expenditure data accounts for a range of subsidies. Firstly, there are several thresholds where the state will reimburse a percentage of expenditure (Appendix A, eTable 2).^21^ These thresholds have increased over recent years.^31,32^ In 2020, the maximum an individual could spend in a year on medicines was DKK 4,190 (€563). Secondly, the data accounts for the subsidies that people with certain disabilities receive,^20^ and the subsidies that municipalities provide toward medicine costs for those on low incomes.^20^ Thirdly, the data accounts for the subsidies a doctor can apply for on behalf of an individual patient for a) a more expensive version of a reimbursed medication or b) a medication that is not on the reimbursed list, if they can outline why it is needed for that specific patient.^33^ The data does not account for the reimbursements associated with private health insurance. Moreover, they do not include medicines provided during in-patient admissions or those provided directly by treatment centres or hospitals (e.g. methadone or chemotherapeutic drugs),^23^ though there is no out-of-pocket charge associated with these medicines. Payment data is exchanged from Danish Kroner to Euro at a rate of 7.5 kroner per euro. The exchange rate was the average annual exchange rate for 2020 published by the European Central Bank.^34^

Also, of note, prescription medicines in Denmark are sold at the same prices in all pharmacies;^35^ this is the price paid by the state plus a uniform markup.^36^ When a medicine is dispensed at the pharmacy, it is required by law that the cheapest alternative be offered, although the patient may opt for a more expensive one,^22^ in which case patients pay the difference in price out-of-pocket. Also, full price has to be paid for certain classes of medications which are not eligible for reimbursement (e.g. benzodiazepines).^23^ All medicines, including those not eligible for general reimbursement such as benzodiazepines, are fully reimbursed for individuals who are terminally ill.^37^

The financial burden of out-of-pocket payments for prescription medicines has been estimated by dividing an adult’s out-of-pocket expenditure on prescription medicines in 2020 by their equivalised household income. Equivalised household income is an adjusted measure of household income using the OECD-modified equivalence scale for household size.^38^ This involves dividing household income by the number of people in one’s household, where a weight of one is applied to the first adult, 0.5 for each additional adult and 0.3 for each child.

To contextualise out-of-pocket prescription medicine expenditure, details of the number of different active repeat prescriptions for long term conditions is included in the results. It was captured on 1^st^ January 2020 and uses a list of ATC codes (eTable 3) which contains most medicines relevant for chronic disease management. It excludes acute items (e.g. most antibiotics and dermatological agents).^23,25^ Though it should be noted that this only applied to the mean number of prescriptions for long-term conditions variable; expenditure analysis includes data for all redeemed prescription medicines, including medicines for both long-term and acute issues.

#### 2.1.3 Covariates

Data on education level, population density, immigration status, age, cohabitation status, gender and household income were captured on 1^st^ January 2020. Age and gender are included as covariates for face validity. Education level, immigration status, population density, household income and cohabitation status were included as covariates because they are potential confounders of the relationship between multimorbidity and out-of-pocket prescription medicine expenditure. For full details of immigration status categories, education categories and population density definitions see eBox 2 and Statistics Denmark.^39,40^

### 2.2 Data analysis

Frequencies and descriptive statistics are used to outline demographics, number of chronic conditions and number of redeemed prescription medicines and to provide an overview of out-of-pocket expenditure on prescription medicines, financial burden, and state subsidies. Descriptive statistics are also used to assess the percentage/number of people spending more than 10% of equivalised household income on prescription medicines. Demographic, medicine and expenditure statistics are stratified by number of chronic conditions.

A two-part hurdle regression model was applied to analyse the relationship between multimorbidity and out-of-pocket prescription medicine expenditure, while adjusting for a range of demographic and socioeconomic variables.^41^ The first part is a probit regression, which models the binary outcome of having any out-of-pocket prescription medicine expenditure. The second part is a generalised linear model (GLM) with log-link, and gamma-distributed errors, which models intensity of out-of-pocket prescription medicine expenditure amongst those with any out-of-pocket prescription medicine expenditure. A GLM of this nature is applied to expenditure data to accommodate the large tail commonly found with this type of data.^42,43^ This two-part hurdle model accommodates the large number of non-spenders, which is often found in out-of-pocket expenditure data.^41^

Unconditional quantile regressions were also conducted to understand the effect of health, demographic and socioeconomic variables on out-of-pocket healthcare expenditure at different points of the expenditure distribution. The quantile regression is presented across nine quantiles to allow for comparison with previous studies of multimorbidity and out-of-pocket healthcare expenditure.^13,44,45^

For the non-quantile regression analysis, two models were fitted, first including variables for face validity: age and sex, and to meet the research aims: number of conditions (details in Appendix A eTable 4). The second model included additional demographics: population density, education, household income, immigrations status and cohabitation status. Household income was divided by one thousand for the relevant models to facilitate interpretable coefficients. Also, as part of a sensitivity analysis, an ordinary least squares (OLS) regression was conducted to assess the relationship between multimorbidity and intensity of out-of-pocket prescription medicine expenditure (Appendix A, eTable 5). To explore whether results differed due to the COVID-19 pandemic beginning in 2020, frequencies and descriptive statistics for 2019 demographics and out-of-pocket prescription medicine expenditure are provided in Appendix A (eTables 6-8), along with regression figures for the hurdle model using 2019 data.

For the unconditional quantile regression, there was one model only, which included the following variables: number of conditions, age, sex, education, cohabitation status, immigration status, population density and household income. Complete-case analysis was used for all regression models in line with previous multimorbidity and healthcare utilisation analysis of Danish healthcare databases.^18^

The non-quantile regression analysis was performed in Stata version 17. The quantile regression was performed in R.

### 2.3 Ethics

Ethical approval and informed consent were not needed as the study was based on de-identified register data encrypted by and stored with Statistics Denmark. The study was approved by the Scientific Board of Statistics Denmark and the Danish Data Protection Agency.

## 3. Results

We identified 5,010,356 adults aged ≥18 years old resident in Denmark in 2020. The average age was 48.5 (SD=18.9) years, 50.3% (N=2,518,879) of the population were female, and 25.8% (N=1,136,309) had 10 years of education or less. Overall, 1,212,033 (24.2%) individuals had multimorbidity (2+ conditions). Individuals with multimorbidity were more likely to be older and have a lower education level. Table 2 provides details of the population characteristics by multimorbidity status.

**Table 1.**
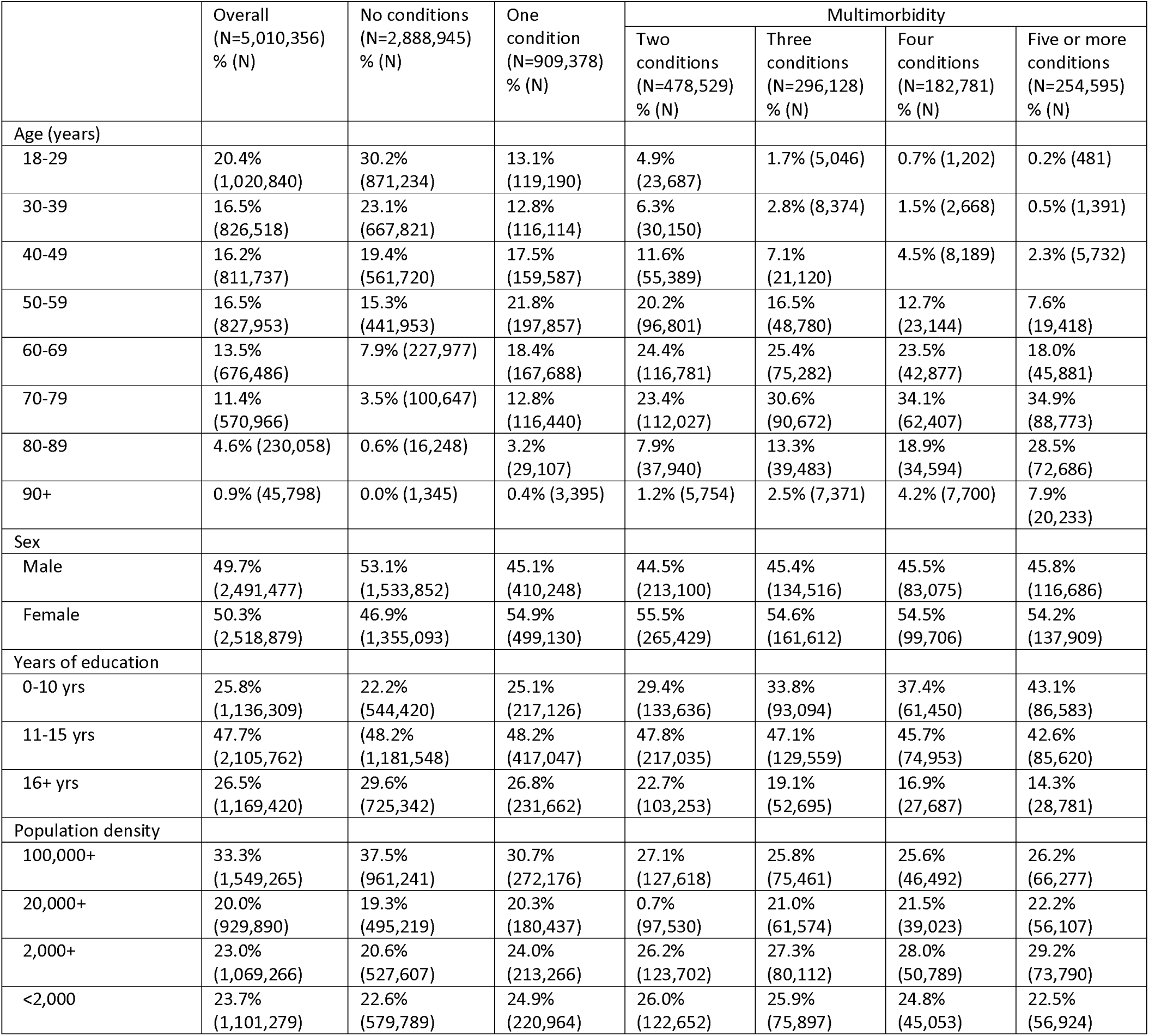
Demographic characteristics of sample.

|  | Overall<br>(N=5,010,356)<br>% (N) | No conditions<br>(N=2,888,945)<br>% (N) | One<br>condition<br>(N=909,378)<br>% (N) | Multimorbidity |  |  |  |
| --- | --- | --- | --- | --- | --- | --- | --- |
|  |  |  |  | Two<br>conditions<br>(N=478,529)<br>% (N) | Three<br>conditions<br>(N=296,128)<br>% (N) | Four<br>conditions<br>(N=182,781)<br>% (N) | Five or more<br>conditions<br>(N=254,595)<br>% (N) |
| Age (years) |  |  |  |  |  |  |  |
| 18-29 | 20.4%<br>(1,020,840) | 30.2%<br>(871,234) | 13.1%<br>(119,190) | 4.9%<br>(23,687) | 1.7% (5,046) | 0.7% (1,202) | 0.2% (481) |
| 30-39 | 16.5%<br>(826,518) | 23.1%<br>(667,821) | 12.8%<br>(116,114) | 6.3%<br>(30,150) | 2.8% (8,374) | 1.5% (2,668) | 0.5% (1,391) |
| 40-49 | 16.2%<br>(811,737) | 19.4%<br>(561,720) | 17.5%<br>(159,587) | 11.6%<br>(55,389) | 7.1%<br>(21,120) | 4.5% (8,189) | 2.3% (5,732) |
| 50-59 | 16.5%<br>(827,953) | 15.3%<br>(441,953) | 21.8%<br>(197,857) | 20.2%<br>(96,801) | 16.5%<br>(48,780) | 12.7%<br>(23,144) | 7.6%<br>(19,418) |
| 60-69 | 13.5%<br>(676,486) | 7.9% (227,977) | 18.4%<br>(167,688) | 24.4%<br>(116,781) | 25.4%<br>(75,282) | 23.5%<br>(42,877) | 18.0%<br>(45,881) |
| 70-79 | 11.4%<br>(570,966) | 3.5% (100,647) | 12.8%<br>(116,440) | 23.4%<br>(112,027) | 30.6%<br>(90,672) | 34.1%<br>(62,407) | 34.9%<br>(88,773) |
| 80-89 | 4.6% (230,058) | 0.6% (16,248) | 3.2%<br>(29,107) | 7.9%<br>(37,940) | 13.3%<br>(39,483) | 18.9%<br>(34,594) | 28.5%<br>(72,686) |
| 90+ | 0.9% (45,798) | 0.0% (1,345) | 0.4% (3,395) | 1.2% (5,754) | 2.5% (7,371) | 4.2% (7,700) | 7.9%<br>(20,233) |
| Sex |  |  |  |  |  |  |  |
| Male | 49.7%<br>(2,491,477) | 53.1%<br>(1,533,852) | 45.1%<br>(410,248) | 44.5%<br>(213,100) | 45.4%<br>(134,516) | 45.5%<br>(83,075) | 45.8%<br>(116,686) |
| Female | 50.3%<br>(2,518,879) | 46.9%<br>(1,355,093) | 54.9%<br>(499,130) | 55.5%<br>(265,429) | 54.6%<br>(161,612) | 54.5%<br>(99,706) | 54.2%<br>(137,909) |
| Years of education |  |  |  |  |  |  |  |
| 0-10 yrs | 25.8%<br>(1,136,309) | 22.2%<br>(544,420) | 25.1%<br>(217,126) | 29.4%<br>(133,636) | 33.8%<br>(93,094) | 37.4%<br>(61,450) | 43.1%<br>(86,583) |
| 11-15 yrs | 47.7%<br>(2,105,762) | 48.2%<br>(1,181,548) | 48.2%<br>(417,047) | 47.8%<br>(217,035) | 47.1%<br>(129,559) | 45.7%<br>(74,953) | 42.6%<br>(85,620) |
| 16+ yrs | 26.5%<br>(1,169,420) | 29.6%<br>(725,342) | 26.8%<br>(231,662) | 22.7%<br>(103,253) | 19.1%<br>(52,695) | 16.9%<br>(27,687) | 14.3%<br>(28,781) |
| Population density |  |  |  |  |  |  |  |
| 100,000+ | 33.3%<br>(1,549,265) | 37.5%<br>(961,241) | 30.7%<br>(272,176) | 27.1%<br>(127,618) | 25.8%<br>(75,461) | 25.6%<br>(46,492) | 26.2%<br>(66,277) |
| 20,000+ | 20.0%<br>(929,890) | 19.3%<br>(495,219) | 20.3%<br>(180,437) | 0.7%<br>(97,530) | 21.0%<br>(61,574) | 21.5%<br>(39,023) | 22.2%<br>(56,107) |
| 2,000+ | 23.0%<br>(1,069,266) | 20.6%<br>(527,607) | 24.0%<br>(213,266) | 26.2%<br>(123,702) | 27.3%<br>(80,112) | 28.0%<br>(50,789) | 29.2%<br>(73,790) |
| <2,000 | 23.7%<br>(1,101,279) | 22.6%<br>(579,789) | 24.9%<br>(220,964) | 26.0%<br>(122,652) | 25.9%<br>(75,897) | 24.8%<br>(45,053) | 22.5%<br>(56,924) |

**Table 2.** Annual out-of-pocket prescription medicine expenditure (€) and financial burden by number of conditions.

|  | Overall | No conditions | One condition | Multimorbidity |  |  |  |
| --- | --- | --- | --- | --- | --- | --- | --- |
|  |  |  |  | Two conditions | Three conditions | Four conditions | Five or more conditions |
| Mean number of medicines for long-term conditions (SD) | 1.5 (2.5) | 0.3 (0.9) | 1.3 (1.8) | 2.7 (2.4) | 4.1 (2.8) | 5.2 (3.1) | 6.9 (3.6) |
| Percentage with any OOP<sup>a</sup> prescription medicine expenditure (N) | 69.8% (3,495,638) | 53.4% (1,541,518) | 85.8% (779,795) | 95.0% (454,467) | 97.8% (289,571) | 98.5% (180,069) | 98.3% (250,218) |
| Total annual OOP prescription medicine expenditure (€) Mean (SD) | 106.01 (169.99) | 43.95 (95.45) | 124.59 (156.08) | 186.60 (188.97) | 236.85 (216.15) | 272.72 (230.23) | 320.46 (275.26) |
| Total annual OOP prescription medicine expenditure (€) Median (IQR) | 38.33 (0.00-153.47) | 7.39 (0.00-47.20) | 80.73 (22.32-179.96) | 149.39 (65.46-256.81) | 196.74 (100.25-322.10) | 234.48 (125.51-373.79) | 274.52 (132.02-441.54) |
| Subsidies (€) Mean (SD) | 166.53 (894.96) | 27.09 (548.43) | 127.91 (758.49) | 265.14 (934.97) | 463.27 (1,397.56) | 654.28 (1,591.67) | 1,005.96 (1,827.27) |
| Equivalised Household Income (€) Mean (SD) | 41,057 (72,802) | 42,374 (59,754) | 42,609 (103,836) | 40,335 (82,842) | 37,334 (58,541) | 34,849 (46,902) | 32,052 (65,355) |
| Financial Burden<sup>b</sup> Mean (SD) | 0.6% (69.4) | 0.4% (73.7) | 0.7% (80.4) | 0.9% (69.7) | 1.1% (32.7) | 1.1% (19.1) | 1.3% (8.8) |
| Percentage with Financial Burden >10% (N) | 0.12% (6,167) | 0.11% (3,266) | 0.14% (1,273) | 0.14% (670) | 0.14% (425) | 0.12% (221) | 0.12% (312) |

<sup>a</sup> The acronym OOP is used for out-of-pocket throughout Table 2
<sup>b</sup> The percentage of equivalised household income spent on prescription medicines in previous 12 months

### 3.1 Multimorbidity, out-of-pocket prescription medicine expenditure and financial burden

The average annual out-of-pocket prescription medicine expenditure for Danish adults was €106.01 (SD=169.99). Those with multimorbidity spent, on average, more than those with zero or one chronic condition. For example, average expenditure for those with two conditions was €186.60 (SD=188.97), whereas for those with one condition, it was €124.59 (SD=156.08), and for those with no conditions, it was €43.95 (SD=95.45). Those with a greater number of conditions had higher average annual out-of-pocket prescription medicine expenditure. Specifically, those with five or more conditions spent an average of €320.46 (SD=275.26) (Fig. 1). Those with multimorbidity also had lower equivalised household income. Specifically, those with five or more conditions had an equivalised household income of €32,052 (SD=65,355) compared to €42,374 (SD=59,754) for those with no conditions and €42,609 (103,836) for those with one condition. This translated into a greater financial burden for those with multimorbidity. With regard to subsidies, the Danish state, on average, covered €166.53 (SD=894.96) annually for prescription medicines for adults in Denmark, which is 61% of overall prescription medicine costs. For those with five or more conditions, this increases to €1005.96 (SD=1827.27), which is 75% of overall prescription medicine costs for that group. For those with one condition, the Danish state, on average, covered €27.09 (SD=548.43) annually for prescription medicines for adults in Denmark, which is 38% of overall prescription medicine costs for that group (Fig. 1). Full details, including details of subsidies, are presented in Table 2. Similar patterns of out-of-pocket prescription medicine expenditure were found for 2019 data (Appendix A eTable 7).

**Figure 1.**
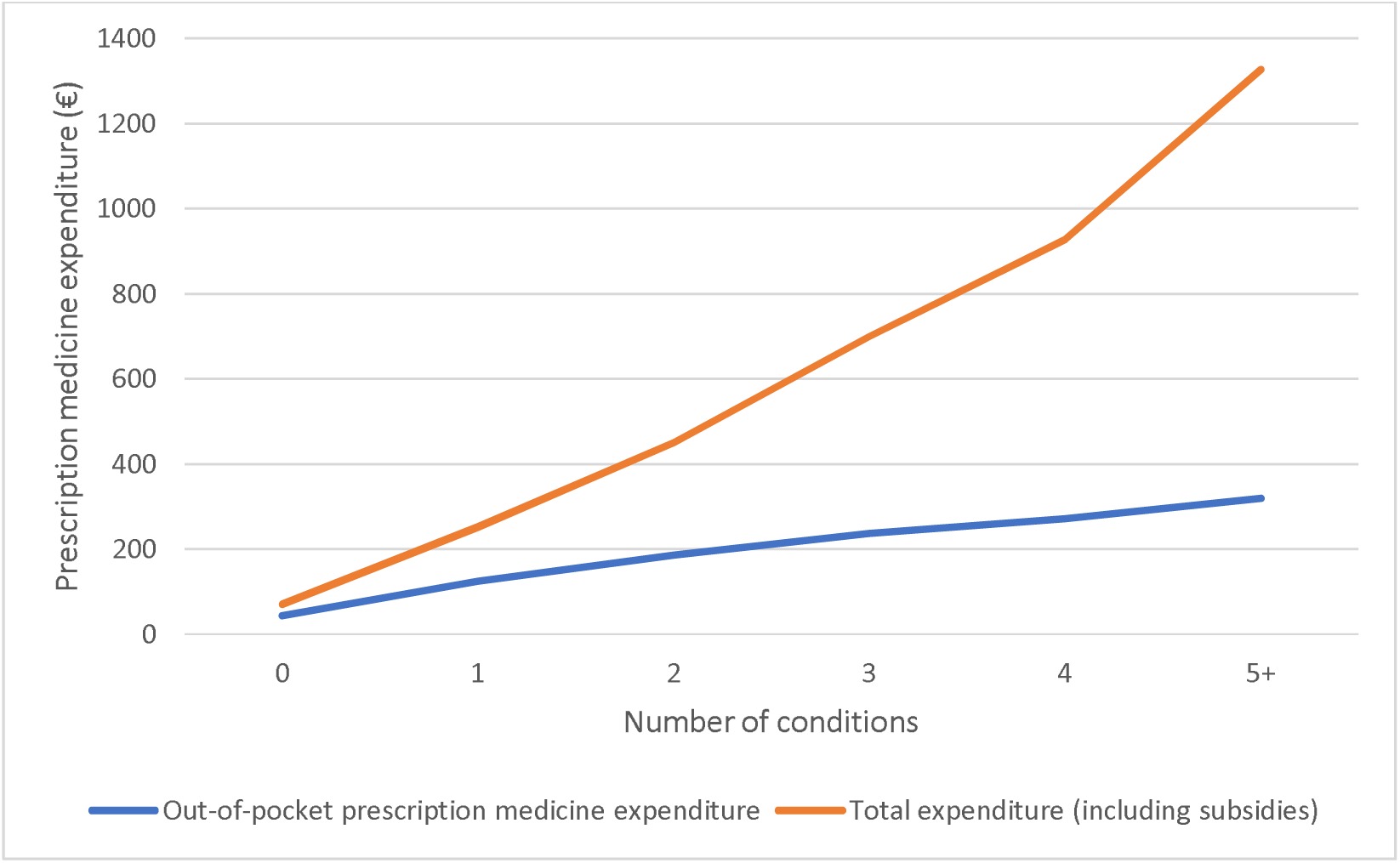
Out-of-pocket prescription medicine expenditure and subsidies by number of conditions

### 3.2 Association between multimorbidity and out-of-pocket prescription medicine expenditure

The probit model (Table 3, part a) revealed that, when demographic and socioeconomic variables were adjusted for, a greater number of conditions was associated with a greater likelihood of any out-of-pocket prescription medicine expenditure. Also, lower odds of any out-of-pocket prescription medicine expenditure was seen in individuals with higher education, those who are not married or cohabiting, those in the lowest income quintile and those who or immigrants or ‘descendants’. Older age was associated with an increased likelihood of any out-of-pocket prescription medicine expenditure, except for those aged 30-49, who were less likely to have any out-of-pocket prescription medicine expenditure than those aged under 30 years.

**Table 3.** Two-part hurdle model examining a) probability of any out-of-pocket prescription medicine expenditure and b) intensity of out-of-pocket prescription medicine expenditure^a^.

| No. of conditions | a) Probit model<br>Odds Ratio<br>(95% confidence interval) | b) GLM<br>Multiplicative difference<br>(95% confidence interval) |
| --- | --- | --- |
| 0 | Ref | Ref |
| 1 | 2.23 (2.23-2.23) | 1.70 (1.70-1.70) |
| 2 | 3.94 (3.94-3.94) | 2.27 (2.26-2.27) |
| 3 | 6.16 (6.15-6.16) | 2.81 (2.81-2.81) |
| 4 | 8.10 (8.09-8.12) | 3.23 (3.23-3.23) |
| 5+ | 9.53 (9.52-9.55) | 3.89 (3.89-3.89) |
| Age group |  |  |
| 18-29 | Ref | Ref |
| 30-39 | 0.99 (0.98-0.98) | 1.10 (1.10-1.10) |
| 40-49 | 0.99 (0.99-0.99) | 1.15 (1.15-1.15) |
| 50-59 | 1.07 (1.07-1.07) | 1.19 (1.19-1.19) |
| 60-69 | 1.20 (1.20-1.20) | 1.20 (1.20-1.20) |
| 70-79 | 1.35 (1.35-1.35) | 1.13 (1.13-1.13) |
| 80-89 | 1.46 (1.46-1.46) | 1.09 (1.09-1.09) |
| 90+ | 1.31 (1.31-1.31) | 1.09 (1.09-1.09) |
| Sex |  |  |
| Male | Ref | Ref |
| Women | 1.62 (1.62-1.62) | 1.10 (1.10-1.10) |
| Education |  |  |
| 0-10 yrs | Ref | Ref |
| 11-15 yrs | 0.97 (0.97-0.97) | 0.96 (0.96-0.96) |
| 16+ yrs | 0.88 (0.88-0.88) | 0.98 (0.98-0.98) |
| Cohabitation status |  |  |
| Single | Ref | Ref |
| Married | 1.08 (1.08-1.08) | 0.95 (0.95-0.95) |
| Cohabiting | 1.06 (1.06-1.06) | 0.92 (0.92-0.92) |
| Immigration status |  |  |
| ≥1 Parent born in Denmark &<br>parent is Danish citizen | Ref | Ref |
| Descendent | 0.84 (0.84-0.84) | 0.86 (0.86-0.86) |
| Immigrant | 0.93 (0.93-0.93) | 0.89 (0.89-0.89) |
| Population density |  |  |
| 100,000+ | Ref | Ref |
| 20,000+ | 1.03 (1.03-1.03) | 1.00 (1.00-1.00) |
| 2,000+ | 1.02 (0.99-1.02) | 1.00 (1.00-1.00) |
| <2,000 | 0.98 (0.98-0.98) | 1.00 (1.00-1.00) |
| Equivalised Income quintile |  |  |
| 1 | Ref | Ref |
| 2 | 1.07 (1.07-1.07) | 1.01 (1.01-1.01) |
| 3 | 1.07 (1.07-1.07) | 1.05 (1.05-1.05) |
| 4 | 1.06 (1.06-1.06) | 1.03 (1.03-1.03) |
| 5 | 1.06 (1.06-1.06) | 1.05 (1.04-1.05) |
| Intercept | 0.99 (0.99-0.99) | 74.74 (74.74-74.74) |

<sup>a</sup>Model b includes only those with any out-of-pocket prescription medicine expenditure
Note: The variables are mutually adjusted.

The GLM (Table 3, part b) found a strong association between number of conditions and intensity of out-of-pocket prescription medicine expenditure amongst those with any out-of-pocket prescription medicine expenditure. Having two chronic conditions was associated with 2.27 times greater out-of-pocket prescription medicine expenditure than those with zero conditions. This translates to €191.80 in expenditure for those with two chronic conditions versus €84.40 for those with no conditions. Having five or more chronic conditions was associated with 3.89 times greater out-of-pocket prescription medicine expenditure than those with zero conditions. This translates into €327.85 in expenditure for those with five or more chronic conditions. Figure 2 shows a graphical representation of the relationship between number of conditions and out-of-pocket prescription medicine expenditure, based on the GLM.

**Figure 2.**
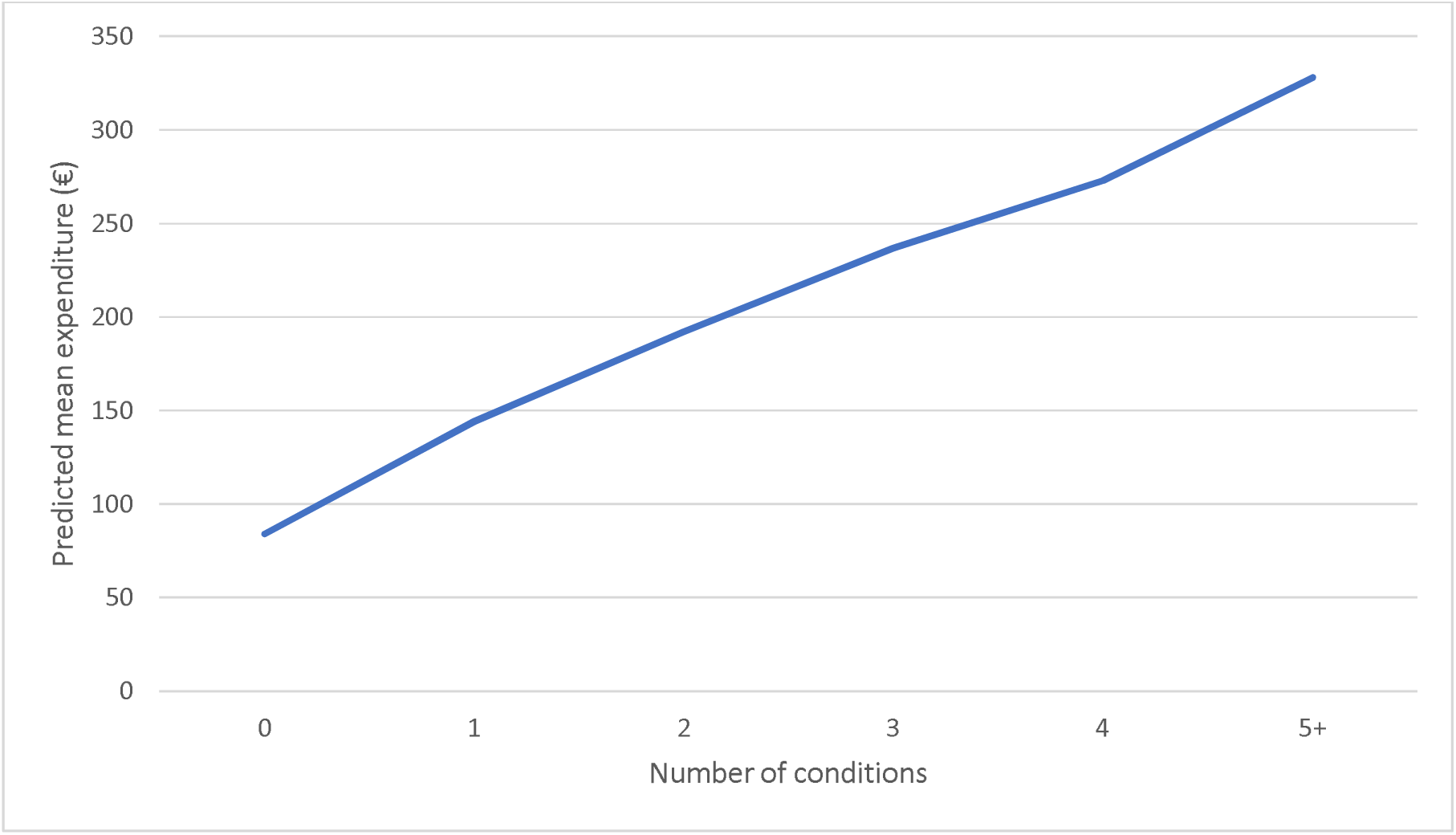
Predicted out-of-pocket prescription medicine expenditure for spenders

Age was also associated with increased out-of-pocket prescription medicine expenditure, peaking amongst those aged 60-69 years whose expenditure was estimated to be 1.20 times greater than those aged 18-29 years. Women were found to have both an increased likelihood of any out-of-pocket prescription medicine expenditure and an increased level of out-of-pocket prescription medicine expenditure. Amongst those incurring any out-of-pocket prescription medicine expenditure, out-of-pocket prescription medicine expenditure for women was estimated to be 1.10 times higher than for men. Higher equivalised household income was associated both with higher likelihood of any out-of-pocket prescription medicine expenditure and with an increased level of out-of-pocket prescription medicine expenditure. Amongst those incurring any out-of-pocket prescription medicine expenditure, out-of-pocket prescription medicine expenditure for those in the highest quintile was estimated to be 1.05 times higher than for the lowest quintile. For both models, similar patterns of results were found when analysing 2019 data (Appendix A eTable 8).

### 3.3 Effect of multimorbidity across the out-of-pocket expenditure distribution

An unconditional quantile regression showed a strong positive association between multimorbidity and out-of-pocket prescription medicine expenditure (Fig. 3). For example, in the group with the lowest expenditure (i.e. quantile 1) those with two conditions spent €26.54 more per annum than those with no conditions, this difference increased to €184.71 in the group with the highest expenditure (i.e. quantile 9). When looking at those with five or more conditions, we see in quantile 1, that this group spent €67.32 more per annum than those with no conditions, whereas, in quantile 9, this group spent €407.88 more than those with no conditions. Details of all coefficients can be seen in Appendix A eTable 9.

**Figure. 3.**
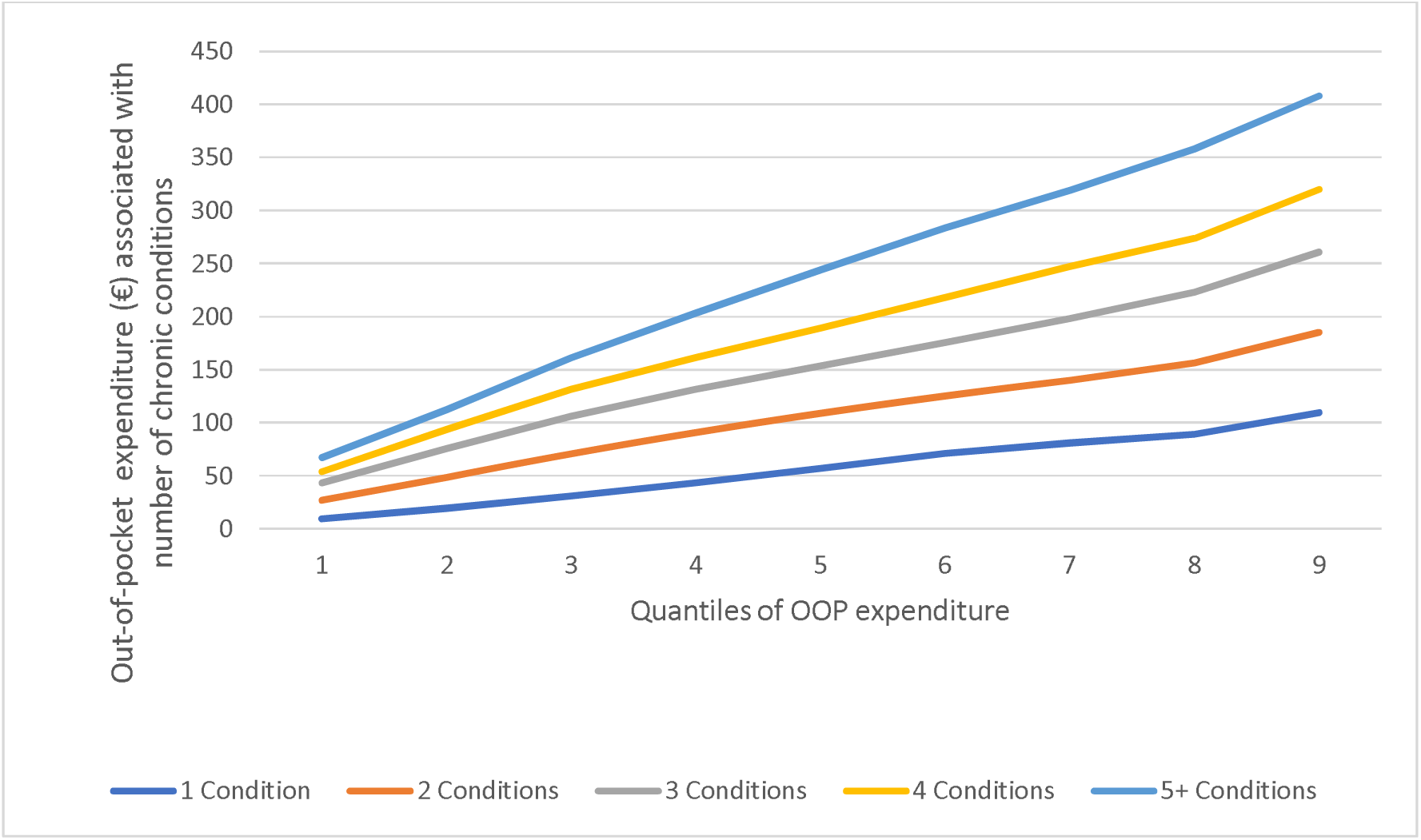
Quantile regression coefficients for effect of number of conditions on out-of-pocket (OOP) prescription medicine expenditure.

## 4. Discussion

For adults in Denmark, having multimorbidity is associated with significantly more out-of-pocket prescription medicine expenditure. In fact, the study found that individuals with multimorbidity, compared to those with zero conditions, incurred 4-7 times more annual out-of-pocket prescription medicine expenditure, depending on their number of chronic conditions. This translated into an average annual out-of-pocket prescription medicine spend of €320 for those with five or more conditions, €187 for those with 2 conditions and €44 for those with no conditions. This relationship was slightly attenuated when controlling for several covariates and analysed among those with any expenditure only. Specifically, our regression models revealed that individuals with multimorbidity incurred 2-4 times more annual expenditure compared to those with zero conditions, independent of demographic and socioeconomic covariates. The quantile regression revealed that those with high expenditure primarily drove this increase. The situation is compounded by the fact that individuals with multimorbidity also tend to have lower household incomes. This means that along with having higher out-of-pocket prescription medicine expenditure, they also have a lower ability to afford these healthcare costs. People with five or more conditions spend 1.3% of their equivalised household income on prescription medicines, whereas people with zero conditions spent 0.4% of their equivalised household income. Though this high expenditure amongst those with more conditions would be far greater if not for the state subsidies on prescription medicine expenditure, whereby those with more prescription medicine expenditure have a larger proportion of their medicine costs paid for by the state.

### 4.1 In the context of other literature

The patterns observed in this study align with previous studies. Specifically, the increased financial burden associated with a greater number of conditions has also been found in other studies.^12–14^ Also, our findings, that those with the highest expenditure primarily drive large out-of-pocket prescription medicine expenditure amongst those with multimorbidity, align with several recent studies examining out-of-pocket healthcare expenditure overall.^13,46,47^ The increase in expenditure associated with multimorbidity is consistent with previous studies; however, it is noteworthy that studies from other countries show a greater increase in out-of-pocket prescription medicine expenditure associated with multimorbidity. A systematic review of out-of-pocket prescription medicine costs found that multimorbidity was associated with five to ten times more out-of-pocket prescription medicine costs, compared to no chronic conditions.^12^ In contrast, our study reveals a four to sevenfold increase (details of absolute expenditure in next paragraph). Though it is important to acknowledge that most of these studies^12^ were conducted in the US where there is limited healthcare coverage^48^ and healthcare expenditure per capita is higher than most other countries.^49^ A previous international review examined both healthcare system costs and out-of-pocket healthcare costs, and it concluded that each additional condition was associated with a ‘near exponential’ increase in healthcare expenditure.^50^ However, our results concerning the relationship between each additional condition and out-of-pocket prescription medicine costs instead showed a near logarithmic relationship, with the percentage increase in costs being just above 50% for each additional condition. The ‘near exponential’ relationship reported previously may be attributable to the preponderance of US studies in the previous systematic review.^50^ Unlike the US, Denmark has a system of universal state subsidies for out-of-pocket prescription medicine expenditure, which is clearly strongly attenuating the relationship between the number of conditions and out-of-pocket prescription medicine expenditure with a near-exponential increase in subsidies associated with each additional condition.

Subsidies in Denmark seem to be keeping out-of-pocket prescription medicine expenditure low when compared with other countries. Using data that is adjusted to 2015 values by Sum and colleagues^12^ and that we have exchanged to Euros using European central bank figures, we find that Danes have lower out-of-pocket prescription medicine expenditure than other countries. For example, in Canada annual out-of-pocket prescription medicine expenditure for people with two or more conditions aged under 65 years was €562 on average and for those aged over 65 years, it was €726.^12^ A US study found that individuals aged 18–64 years with four or more conditions spent, on average, €1,032 annually.^12^ Only Korea reported lower expenditure rates than Denmark, where individuals aged 20 years or over with three or more conditions spent on average €234 annually on prescription medicines.^12^ It is important to note that these figures would likely increase when adjusted for inflation to 2020 values. With regard to European examples, an analysis of 2016 data in Ireland found that individuals with three or more conditions spent €453.5 annually on medicines.^13^ However, it should be noted that the Irish analysis only included those with any expenditure and analysis was limited to those aged 50 years and over.^13^

Despite these comparisons showing relatively low out-of-pocket prescription medicine expenditure for people with multimorbidity in Denmark, a 2021 analysis of countries in the European Union revealed that the percentage of outpatient medicines expenditure covered by the Danish state is 43%, which is much lower than the EU average of 57%.^51^ This inconsistency might be explained by overall expenditure on pharmaceuticals and medical devices in Denmark being much lower than the EU average.^51^ Several factors contribute to this, including the legal mandate in Danish community pharmacies, when filling a prescription, to substitute in the cheapest generic equivalent of the prescribed medicine.^52^ Also, according to the Association of Danish Pharmacists, Denmark has the lowest prices for generic medicines in Europe^53^ likely due to its medication tendering system, which is considered one of the most competitive and flexible systems in Europe.^54^

It is likely that expenditure amongst Danish individuals with multimorbidity would be higher if the population were fully adherent to their medication. In 2019, 6% reported an unmet prescription medicine need due to financial reasons.^55^ A systematic review of multimorbidity and out-of-pocket prescription medicine expenditure found that cost-related non-adherence was a common coping strategy for out-of-pocket prescription medicine costs.^12^ Non-adherence can have a range of negative health outcomes such as disease progression and hospitalisation.^56,57^ The association between increased income and increased expenditure in our results potentially indicates that those with lower incomes are engaging in cost-related non-adherence. However, it’s notable that the difference in out-of-pocket prescription medicine expenditure in the GLM is not very high with the highest income quintile spending 1.05 times more than the lowest income quintile. This may be attributable to the low-levels of income inequality in Denmark.^58^

### 4.1 Implications

The substantial difference between the percentage of outpatient medicines expenditure covered by the Danish state compared to the EU average,^51^ suggests potential capacity for expanding medication entitlements. Consideration should be given to expanding free access to low-income groups, as they are both more likely to have multimorbidity^19^ and less likely to afford their medicines.^12^

Additionally, consideration could also be given to greater entitlements for certain patient populations. Free antipsychotic medications for those with schizophrenia for the first two years after diagnosis was introduced by the Danish state in 2008.^59^ Consideration could be given to widening the patient group and list of medications covered by this scheme. Individuals with severe mental illnesses in Denmark are more likely of lower socioeconomic status,^60^ and these conditions are associated with an increased risk of developing physical multimorbidity.^17,61^. Therefore, free prescription medicines for individuals with severe mental disorders could also reduce the high out-of-pocket prescription medicine expenditure for many with multimorbidity and low incomes.

In 2022, a new contract with GPs was introduced in Denmark.^62^ Within this contract, a pilot initiative was included for patients with multimorbidity to extend consultation time to facilitate better medication overview and time for GPs to better coordinate with other healthcare professionals.^63^ The initiative can potentially reduce prescribing, particularly potentially inappropriate prescribing, for individuals with multimorbidity.^64^ Longer consultation times could also provide opportunities for cost-of-care conversations whereby a healthcare professional discusses out-of-pocket healthcare costs and health coverage.^65^ Such conversations have the potential to reduce costs to the patient,^66^ increase their adherence^67^ and reduce social inequality in medical treatment.^68^

Another consideration for optimising prescribing practice and subsequently potentially reducing costs is an increase in the provision of medication reviews. Medication reviews involve a healthcare worker, typically a doctor or pharmacist, assessing the individual’s medicine regimen to identify medication-related issues and suggesting strategies for addressing these issues.^69^ A 2017 overview of systematic reviews^69^ found that medication reviews significantly reduce the number of medications a person is prescribed and consequently medication costs. The capital region of Denmark has a program where GPs are employed part-time to visit other GPs in the region to support rational prescribing;^70^ expanding this program could be considered. Alternatively, consideration could be given to integrating a pharmacist into a general practice setting. The pharmacist would fulfill several duties including medication reviews. This has been implemented in the UK and is being trialled in Ireland.^71^ This GP-pharmacist model was trialed in Denmark in the early 2000s but it was not implemented.^72^ Though forms of medication reviews by community pharmacies are already reimbursed in Denmark.^73^

Future research could explore the relationship between out-of-pocket expenditure for aspects of care other than prescription medicines, as these account for only about 36% of out-of-pocket healthcare expenditure in Denmark.^74^ Future research in the area of medicines, out-of-pocket expenditure and multimorbidity could involve a more detailed analysis of the different entitlements available in Denmark. This could include an effort to model a range of changes to the entitlement system and how this would affect different groups. Also, given that those with the highest out-of-pocket prescription medicine expenditure are largely driving the increase in costs for people with multimorbidity, research focusing on this group’s out-of-pocket expenditure would be valuable. Specifically, researching drivers of the expenditure and possible interventions to reduce it. Additionally, investigating the travel costs associated with medicines and accessing healthcare, considering factors such as rurality and type of transport, should be considered in future research.

### 4.2 Strengths & Limitations

The inclusion of almost the entire Danish adult population, is a major strength of the current study, thereby increasing the representation of groups often underrepresented in survey research (e.g. minority ethnic groups^75^ or those with a cognitive impairment^76^) which is commonly used to assess out-of-pocket healthcare expenditure.^12–15^ Though it is likely that certain groups, such as undocumented migrants, were not captured by the database. Another strength of these national databases is that they provide direct measures of expenditure which are more reliable than self-report data. Self-report data is commonly used in similar studies.^11,13,14^ Unlike most studies in this area, that only focus on older adults,^11,13,14^ our study included all adults aged 18 years and over. Furthermore, we conducted a detailed analysis of subsidies for prescription medicine, which most studies in this area do not provide.^11^ A further strength is the use of a more extensive list of conditions for multimorbidity, compared to other similar studies.^11^ Use of the Danish Multimorbidity Index Algorithm is also a strength; it has been shown to align with national and international prevalence estimates for chronic conditions with a large public health impact (e.g. mental health conditions, diabetes, COPD, heart disease and cancer).^77^ It has also shown face validity based on review by national and international experts.^77^

There are some limitations. Our study did not examine the travel costs or other opportunity costs associated with purchasing prescription medicines. Also, the 2020 data may have been influenced by distinct variations arising from the onset of the COVID-19 pandemic.^78^ However, an analysis of 2019 data (Appendix A) revealed similar patterns of demographics and out-of-pocket prescription medicine expenditure. Finally, private health insurance can cover part or all of prescription costs,^20,79^ our analysis did not account for the cost of private health insurance to individuals or the reimbursements they may receive as part of their private health insurance. Approximately 42% of people in Denmark purchase private health insurance.^20^ If private health insurance reimbursements for prescription medicines were included in the analysis this would show lower out-of-pocket prescription medicine expenditure overall. However, this reduction is likely to be unequally distributed and benefit those with fewer chronic conditions and higher socioeconomic status most.^80^

### 4.3 Conclusion

Multimorbidity is associated with increased out-of-pocket prescription medicine expenditure in Denmark, particularly impacting those with lower average incomes. Nevertheless, the Danish entitlements system reduces this financial burden. Overall, these patterns are similar to those observed internationally and are likely contributing to cost-related non-adherence. To address these issues, policymakers should consider expanding entitlements, increasing the provision of medication reviews, and encouraging cost-of-care conversations.

## Data Availability

The dataset is available at Statistics Denmark (https://www. dst.dk/en) and the Danish Health Data Authority (https://sundhedsdatastyrelsen.dk/da/english/). Data access is restricted to authorized research institutions by Danish law.

## Author contributions

JL was involved in the conception, design, analysis and reporting of the study. CHV and APr conducted the analysis for the study. SMS was involved in the conception, design and reporting of the study. TSV, APa and LDC were involved in the reporting of the study. APr was involved in the conception, design, and reporting of the work. JL, CHV, SMS, TSV, LDC, APa and APr have all read and approved the final manuscript and agree to be accountable for all aspects of the work.

