## Appendix A for "The Association between Multimorbidity and Out-of-Pocket Expenditures for Prescription Medicines among Adults in Denmark: A Population-Based Register Study"

| eTable 1. Danish Multimorbidity Index coding definitions | | | | | | | |
| --- | --- | --- | --- | --- | --- | --- | --- |
| **Category** | **Disease group** | **Origin^a^** | **Coding definition^b^** | **Diagnosis codes (ICD-10)** | **Diagnosis time frame** | **Drug codes (ATC)** | **Prescription time frame** |
| **Circulatory system** | Hypertension | a, b | Diagnosis AND/OR prescriptions of antihypertensives, if not ischemic heart disease or heart failure (or kidney disease: only diuretics) | I10-I13, I15 | Ever | C02, C04, C07, C08, C09, C03 | Twice last year |
|  | Dyslipidaemia | b | Diagnosis AND/OR drug prescription for lipid-lowering drugs if not ischemic heart disease. | E78 | Last two years | C10 | Twice last year |
|  | Ischemic heart disease | a, b | Diagnosis AND/OR prescription for antianginal drug | I20-I25 | Ever | C01DA | Twice last year |
|  | Atrial fibrillation | a, b | Diagnosis | I48 | Ever |  |  |
|  | Heart failure | a, b | Diagnosis | I50 | Ever |  |  |
|  | Peripheral artery occlusive disease | a, b | Diagnosis | I70-I74 | Ever |  |  |
|  | Stroke | a, b | Diagnosis | I60-I64, I69 | Ever |  |  |
| **Endocrine system** | Diabetes mellitus | a, b | Diagnosis AND/OR prescription of antidiabetics | E10-E14 | Ever | A10A, A10B | Twice last year |
|  | Thyroid disorder | a, b | Diagnosis AND/OR prescription of thyroid therapy drugs | E00-E05, E061-E069, E07 | Last two years | H03 | Twice last year |
|  | Gout | b | Diagnosis | E79, M10 | Ever |  |  |
| **Pulmonary system and allergy** | Chronic pulmonary disease | a, b | Prescription for obstructive airway disease drugs |  |  | R03 | Twice last year |
|  | Allergy | b | Prescription for non-sedative antihistamines AND/OR nasal antiallergics |  |  | R06AX, R06AE07, R06AE09, R01AC, R01AD | Twice last year |
| **Gastrointestinal system** | Ulcer/chronic gastritis | a, b | Diagnosis | K221, K25-K28, K293-K295 | Ever |  |  |
|  | Chronic liver disease | a, b | Diagnosis | B16-B19, K70-K74, K766, I85 | Ever |  |  |
|  | Inflammatory bowel disease | a, b | Diagnosis | K50-K51 | Ever |  |  |
|  | Diverticular disease of intestine | a, b | Diagnosis | K57 | Ever |  |  |
| **Urogenital system** | Chronic kidney disease | a, b | Diagnosis | N03, N11, N18-N19 | Ever |  |  |
|  | Prostate disorders | a, b | Diagnosis AND/OR prescription of prostate hyperplasia therapy drugs | N40 | Ever | C02CA, G04C | Twice last year |
| **Musculoskeletal system** | Connective tissue disorders | a, b | Diagnosis | M05-M06, M08-M09, M30-M36, D86 | Ever |  |  |
|  | Osteoporosis | b | Diagnosis AND/OR prescription for osteoporosis drugs | M80-M82 | Ever | M05B, G03XC01, H05AA | Twice last year |
|  | Painful condition | a, b | Repeated prescriptions of analgesics |  |  | N02A, N02BA51,N02BE, M01A, M02A | Four times last year |
| **Danish Multimorbidity Index coding definitions (cont.)** | | | | | | | |
| **Category** | **Disease group** | **Origin^a^** | **Coding definition^b^** | **Diagnosis codes (ICD-10)** | **Diagnosis time frame** | **Drug codes (ATC)** | **Prescription time frame** |
| **Hematological system** | HIV/AIDS | b | Diagnosis | B20-B24 | Ever |  |  |
|  | Anaemias | b | Diagnosis | D50-D53, D55-D59, D60-D61, D63-D64 | Last two years |  |  |
| **Cancers** | Cancer | a, b | Diagnosis | C00-C43, C45-C97 | Last five years |  |  |
| **Neurological system** | Vision problem | a, b | Diagnosis | H40, H25, H54 | Ever |  |  |
|  | Hearing problem | a, b | Diagnosis | H90-H91, H931 | Ever |  |  |
|  | Migraine | a, b | Diagnosis AND/OR prescription of specific anti-migraine drugs | G43 | Last two years | N02C | Twice last year |
|  | Epilepsy | a, b | Diagnosis AND prescription of anti-epileptics | G40-G47 | Ever | N03 | Twice last year |
|  | Parkinson's disease | a, b | Diagnosis | G20-G22 | Ever |  |  |
|  | Multiple sclerosis | a | Diagnosis | G35 | Ever |  |  |
|  | Neuropathies | b | Diagnosis | G50-G64 | Last two years |  |  |
| **Mental health conditions** | Mood, stress-related, or anxiety disorders | a, b | Diagnosis | F32-F34, F40-F48 | Last two years |  |  |
|  | Psychological distress | a, b | Prescription of antidepressants if not other mental disorder |  |  | N06A | Twice last year |
|  | Alcohol problems | a, b | Diagnosis | F101-F109 | Last two years |  |  |
|  | Substance abuse | a, b | Diagnosis | F11-F16, F18-F19 | Last two years |  |  |
|  | Anorexia/bulimia | a | Diagnosis | F50 | Last two years |  |  |
|  | Bipolar affective disorder | a, b | Diagnosis AND/OR prescription of lithium salts | F30-F31 | Ever | N05AN | Twice last year |
|  | Schizophrenia or schizoaffective disorder | a, b | Diagnosis | F20, F25 | Ever |  |  |
|  | Dementia | a, b | Diagnosis AND/OR prescription of anti-dementia drugs | F00-F03, F051, G30 | Ever | N06D | Twice last year |
| Abbreviations: ICD-10, International Classification of Diseases, version 10; ATC, Anatomical Therapeutic Chemical Classification System; HIV, human immunodeficiency virus; AIDS, acquired immunodeficiency syndrome.  ^a^ICD-10 diagnosis code recorded or redeemed prescription of ATC-coded drug registered within defined time frames in national health registers (the Danish National Patient Register and the Danish National Prescription Registry.)  ^b^a: Barnett, b: Other index (Van den Bussche, Huber, Charlson, and/or Elixhauser) | | | | | | | |

### eTable 2. annual reimbursement thresholds for prescription medicines for adults in Denmark (2020)

| **Annual personal expenditure on reimbursable medicine before deduction of reimbursement*** | **Reimbursement** |
| --- | --- |
| DKK 0-995 (€0-134) | 0% |
| DKK 995-1,655 (€134-222) | 50% |
| DKK 1,655-3,590 (€222- 482) | 75% |
| In excess of DKK 3,590 (€482) | 85% |
| In excess of DKK 19,465 (€2,615) (patient's co-payment= DKK 4,190 (€563)) | 100% |

*patients pay the full difference if they go for a branded as opposed to a generic medicine

### eTable 3. ATC codes used to analysed number of different active prescriptions

| A02AA0 | A02AB | A02BA | A02BC | A03AA | A03AB | A03B | A03FA0 | A04AD0 | A06A |
| --- | --- | --- | --- | --- | --- | --- | --- | --- | --- |
| A06AC | A07EC0 | A10A | A10BA0 | A10BB0 | A10BB1 | A10BD0 | A10BD1 | A10BD2 | A10BG0 |
| A10BH0 | A10BJ0 | A10BK0 | A10BX0 | A11CC | A12AA | A12AX | B01AA | B01AC0 | B01AC2 |
| B01AC3 | B01AE | B01AF | B03AB | B03BB | C01AA0 | C01BD0 | C01DA | C02 | C02AB |
| C0C02CA | C03 | C03A | C03AA | C03BA | C03C | C03D | C03DA | C03EB | C04 |
| C07AA0 | C07AB0 | C07AB1 | C07AG | C07AG0 | C07BB0 | C07CB0 | C08 | C08DA0 | C08DB0 |
| C09 | C09A | C09B | C09C | C09D | C09DX0 | C10 | C10AA | C10B | G02BB |
| G03AA0 | G03AA1 | G03AB0 | G03AC0 | G03AC1 | G03AD0 | G03BA0 | G03CA0 | G03CC0 | G03CX0 |
| G03DA0 | G03DB0 | G03FA0 | G03FA1 | G03FB0 | G03GA0 | G03GA1 | G03GA3 | G03HA0 | G03HB0 |
| G03XB0 | G03XC0 | G03XX0 | G04BD0 | G04BD1 | G04BE0 | G04BE1 | G04BE3 | G04BX1 | G04C |
| G04CA | G04CB | H02AB0 | H03 | H05AA | J07AL | J07BB | L04AA0 | L04AA1 | L04AA2 |
| L04AA3 | L04AB0 | L04AC0 | L04AD0 | L04AX0 | M01AA0 | M01AB0 | M01AB1 | M01AB5 | M01AC0 |
| M01AE0 | M01AE1 | M01AE5 | M01AG0 | M01AH0 | M01AX0 | M02A | M03BX0 | M04AA | M04AC0 |
| M05B | M05BA0 | M05BB0 | N02A | N02AA0 | N02AA5 | N02AB0 | N02AE0 | N02AG0 | N02AJ0 |
| N02AX0 | N02BA | N02BA0 | N02BA5 | N02BA7 | N02BB5 | N02BE | N02BE0 | N02BE5 | N02BG1 |
| N02CA5 | N02CA7 | N02CC0 | N02CD0 | N02CX0 | N03AA | N03AA0 | N03AB0 | N03AD0 | N03AE0 |
| N03AF0 | N03AG0 | N03AX0 | N03AX1 | N03AX2 | N04A | N04B | N04BC0 | N05AA0 | N05AB0 |
| N05AC0 | N05AD0 | N05AE0 | N05AF0 | N05AG0 | N05AH0 | N05AL0 | N05AN0 | N05AX0 | N05AX1 |
| N05BA0 | N05BA1 | N05BB0 | N05BE0 | N05CC0 | N05CD0 | N05CF0 | N05CH0 | N06AA0 | N06AA1 |
| N06AA2 | N06AB0 | N06AB1 | N06AF0 | N06AG0 | N06AX0 | N06AX1 | N06AX2 | N06BA0 | N06BA1 |
| N06BC0 | N06DA0 | N06DX0 | N07AA0 | N07AA5 | N07BA0 | N07BB0 | N07BC0 | N07BC5 | N07CA0 |
| N07XX0 | N07XX1 | P01BA | R01AC | R01AD | R01AX0 | R03AC0 | R03AC1 | R03AK0 | R03AK1 |
| R03AL0 | R03AL1 | R03BA0 | R03BB0 | R03CA0 | R03CC0 | R03CC1 | R03DA0 | R03DC0 | R03DX0 |
| R05DA0 | R06AA0 | R06AA1 | R06AD0 | R06AE0 | R06AX1 | R06AX2 | S01ED | S01EE |  |

### eBox 1. Calculation process for those residing in Denmark for less than 12 months or having died in the given year

For those residing in Denmark for less than 12 months or having died in the given year, their expenditure has been extrapolated to estimate expenditure for 12 months. For example, if an individual only resided in Denmark for six months, then their expenditure will be multiplied by two and then reimbursements will be applied.

### eBox 2. Definitions of immigration status, education status and population density

Immigration status is composed of three categories; those who, regardless of place of birth, have at least one parent who is both a Danish citizen and born in Denmark (parent born in Denmark & parent is Danish citizen), born outside of Denmark to parents born outside Denmark (immigrant) and born in Denmark to parents born outside Denmark (descendant).

Education is categorised into three levels 0-10 years, 11-15 years and 16+ years. There three categories correspond to *below upper secondary, upper secondary,* and *tertiary* education levels, respectively.^1^

Population density is defined as the number of people living in a defined area. For full definitions of immigration status and population density see Statistics Denmark.^2,3^

### eTable 4. Two-part hurdle model only including variables for face validity

| No. of conditions | Probit  Odds Ratio  (95% confidence interval) | GLM  Multiplicative difference  (95% confidence interval) |
| --- | --- | --- |
| 0 | Ref | Ref |
| 1 | 2.39 (2.39-2.39) | 1.72 (1.72-1.72) |
| 2 | 3.97 (3.97-3.97) | 2.30 (2.30-2.30) |
| 3 | 5.64 (5.64-5.64) | 2.83 (2.83-2.83) |
| 4 | 6.82 (6.81-6.83) | 3.25 (3.25-3.25) |
| 5+ | 7.32 (7.31-7.32) | 3.91 (3.91-3.91) |
| Age group |  |  |
| 18-29 | Ref | Ref |
| 30-39 | 0.87 (0.87-0.87) | 1.08 (1.08-1.08) |
| 40-49 | 1.06 (1.06-1.06) | 1.14 (1.14-1.14) |
| 50-59 | 1.22 (1.13-1.13) | 1.19 (1.19-1.19) |
| 60-69 | 1.39 (1.39-1.39) | 1.21 (1.21-1.21) |
| 70-79 | 1.55 (1.55-1.55) | 1.14 (1.14-1.14) |
| 80-89 | 1.53 (1.53-1.53) | 1.12 (1.12-1.12) |
| 90+ | 1.19 (1.19-1.19) | 1.15 (1.15-1.15) |
| Sex |  |  |
| Male | Ref | Ref |
| Women | 1.55 (1.55-1.55) | 1.1 (1.10-1.10) |
| Intercept | 0.84 (0.84-0.84) | 70.67 (70.6-70.67) |

### eTable 5. OLS model examining and intensity of OOP prescription medicine expenditure amongst spenders only

| No. of conditions | Coefficient (95% confidence interval) |
| --- | --- |
| 0 | Ref |
| 1 | 0.666 (0.6656,0.6659) |
| 2 | 1.063 (1.0632,1.0636) |
| 3 | 1.333 (1.3326,1.3331) |
| 4 | 1.500 (1.499-1.500) |
| 5+ | 1.700 (1.6996,1.7002) |
| Age group |  |
| 18-29 | Ref |
| 30-39 | 0.010 (0.0102,0.0107) |
| 40-49 | 0.076 (0.0756,0.0761) |
| 50-59 | 0.157 (0.1567,0.1571) |
| 60-69 | 0.191 (0.1910,0.1915) |
| 70-79 | 0.095 (0.0943,0.0949) |
| 80-89 | 0.062 (0.0620,0.0627) |
| 90+ | 0.069 (0.0678,0.0692) |
| Sex |  |
| Male | Ref |
| Women | 0.151 (0.1505,0.1507) |
| Education |  |
| 0-10 yrs | Ref |
| 11-15 yrs | 0.001 (0.0009,0.0012) |
| 16+ yrs | 0.004 (0.0043,0.0046) |
| Cohabitation status |  |
| Single | Ref |
| Married | -0.019 (-0.0193,-0.0189) |
| Cohabitating | -0.071 (-0.0707,-0.0703) |
| Immigration status |  |
| Danish born, Danish parents | Ref |
| Descendent | -0.151 (-0.0193,-0.0189) |
| Immigrant | -0.102 (-0.0707,-0.0703) |
| Population density |  |
| 100,000+ | Ref |
| 20,000+ | 0.003 (0.0032,0.0036) |
| 2,000+ | 0.005 (0.0048,0.0052) |
| <2,000 | -0.003 (-0.0035,-0.0032) |
| Income quintile |  |
| 1 | Ref |
| 2 | 0.024 (0.0239,0.0243) |
| 3 | 0.123 (0.1230,0.1234) |
| 4 | 0.092 (0.0917,0.0921) |
| 5 | 0.088 (0.0873,0.0877) |
| Intercept | 3.590 (3.5893,3.5898) |

### eTable 6. Demographic characteristics of sample (2019)

|  | Overall (N=4,950,034)  % (N) | No conditions (N=2,817,603)  % (N) | One condition (N=902,102)  % (N) | Multimorbidity | | | |
| --- | --- | --- | --- | --- | --- | --- | --- |
|  |  |  |  | Two conditions (N=479,580)  % (N) | Three conditions (N=299,814)  % (N) | Four conditions (N=187,019)  % (N) | Five or more conditions (N=263,916)  % (N) |
| Age (years) |  |  |  |  |  |  |  |
| 18-29 | 20.6 (1,018,338) | 30.9  (869,947) | 13.1 (118,510) | 4.9  (23,388) | 1.6  (4,833) | 0.6  (1,181) | 0.2  (479) |
| 30-39 | 16.2  (801,207) | 22.8  (642,653) | 12.8 (115,457) | 6.3  (30,279) | 2.9  (8,627) | 1.4  (2,694) | 0.6  (1,497) |
| 40-49 | 16.4  (809,870) | 19.6  (552,812) | 18.0 (162,472) | 12.0  (57,649) | 7.4  (22,123) | 4.7  (8,743) | 2.3  (6,071) |
| 50-59 | 16.6  (821,540) | 15.2  (427,820) | 22.0 (198,799) | 20.7  (99,315) | 16.9  (50,666) | 13.1  (24,513) | 7.7  (20427) |
| 60-69 | 13.6  (674,178) | 7.7  (217,096) | 18.5 (166,507) | 24.8 (118,893) | 26.1  (78,291) | 24.0  (44,873) | 18.4  (48,518) |
| 70-79 | 11.3  (558,171) | 3.3  (92,328) | 12.3 (111,141) | 22.8 (109,272) | 30.1  (90,308) | 33.9  (63,372) | 34.8  (91,750) |
| 80-89 | 4.5 (221,358) | 0.5 (13,763) | 2.9 (26,105) | 7.4 (35,307) | 12.6 (37,798) | 18.1 (33,823) | 28.3 (74,562) |
| 90+ | 0.9 (45,372) | 0.0 (1,184) | 0.3 (3,111) | 1.1 (5,477) | 2.4 (7,168) | 4.2 (7,820) | 7.8 (20,612) |
| Sex |  |  |  |  |  |  |  |
| Male | 49.7 (2,459,841) | 53.1 (1,496,508) | 45.1 (406,769) | 44.6 (213,919) | 45.6 (136,660) | 45.5  (85,039) | 45.8 (120,946) |
| Female | 50.3 (2,490,193) | 46.9 (1,321,095) | 54.9 (495,333) | 55.4 (265,661) | 54.4 (163,154) | 54.5 (101,980) | 54.2 (142,970) |
| Years of education |  |  |  |  |  |  |  |
| 0-10 yrs | 82.2 (4,068,328) | 75.8 (2,135,482) | 88.7 (800,568) | 90.9 (436,148) | 92.0 (275,815) | 92.6 (173,243) | 93.6 (247,072) |
| 11-15 yrs | 16.1 (798,816) | 21.9 (615,938) | 10.0 (89,828) | 8.4 (40,347) | 7.7 (23,009) | 7.1 (13,336) | 6.2 (16,358) |
| 16+ yrs | 1.7 (82,890) | 2.3 (66,183) | 1.3 (11,706) | 0.6 (3,085) | 0.3 (990) | 0.2 (440) | 0.2 (486) |
| Population density |  |  |  |  |  |  |  |
| 100,000+ | 33.3 (1,553,652) | 37.6  (960,631) | 30.6 (270,888) | 27.0 (128,277) | 25.8  (76,855) | 25.7  (47,845) | 26.3  (69,156) |
| 20,000+ | 19.6  (915,212) | 18.9  (484,369) | 19.9 (176,282) | 20.3  (96,423) | 20.6  (61,201) | 21.2  (39,447) | 21.9  (57,490) |
| 2,000+ | 23.3 (1,086,967) | 20.8  (532,419) | 24.5 (216,514) | 26.5 (125,806) | 27.6  (82,258) | 28.3  (52,725) | 29.4  (77,245) |
| <2,000 | 23.7 (1,107,488) | 22.7  (579,247) | 25.0 (221,816) | 26.1 (123,944) | 26.0  (77,295) | 24.7  (46,025) | 22.5  (59,161) |

### eTable 7. Annual out-of-pocket (OOP) prescription medicine expenditure (€) and household income by number of conditions (2019)

|  | Overall | No conditions | One condition | Multimorbidity | | | |
| --- | --- | --- | --- | --- | --- | --- | --- |
|  |  |  |  | Two conditions | Three conditions | Four conditions | Five or more conditions |
| Mean number of medicines (SD) | 1.5 (2.5) | 0.3 (0.9) | 1.4 (1.8) | 2.8 (2.5) | 4.1 (2.9) | 5.1 (3.3) | 6.2 (4.0) |
| Percentage with any OOP prescription medicine expenditure (N) | 70.9% (3,511,136) | 54.6% (1,537,724) | 86.3% (778,752) | 95.3% (457,144) | 97.9% (293,536) | 98.6% (184,359) | 98.4% (259,621) |
| Total annual OOP prescription medicine expenditure^a^ (€)  Mean (SD) | 105.58 (169.02) | 184.463 (184.6888) | 236.1949 (215.3553) | 184.463 (184.6888) | 236.1949 (215.3553) | 272.1376 (228.6279) | 322.78 (278.04) |
| Equivalised Household Income (€)  Mean (SD) | 39,377 (130,000) | 40,515 (128,480) | 41,065 (206,537) | 38,780 (50,301) | 36,295 (82,402) | 33,848 (50,442) | 30,991 (42,022) |

### eTable 8. Two-part hurdle model examining a) probability of any out-of-pocket prescription medicine expenditure and b) intensity of OOP prescription medicine expenditure (2019)

| No. of conditions | a) Probit model  Odds ratio  (95% confidence interval) | b) GLM  Multiplicative change  (95% confidence interval) |
| --- | --- | --- |
| 0 | Ref | Ref |
| 1 | 2.25 (2.25-2.25) | 1.75 (1.75-1.75) |
| 2 | 4.06 (4.06-4.06) | 2.38 (2.38-2.38) |
| 3 | 6.27 (6.27-6.27) | 2.97 (2.97-2.97) |
| 4 | 8.22 (8.21-8.22) | 3.42 (3.42-3.43) |
| 5+ | 9.24 (9.23-9.26) | 4.18 (4.18-4.18) |
| Age group |  |  |
| 18-29 | Ref | Ref |
| 30-39 | 0.99 (0.99-0.99) | 1.10 (1.10-1.10) |
| 40-49 | 0.97 (0.98-0.97) | 1.15 (1.15-1.15) |
| 50-59 | 1.04 (1.04-1.04) | 1.19 (1.19-1.20) |
| 60-69 | 1.15 (1.15-1.15) | 1.23 (1.23-1.23) |
| 70-79 | 1.28 (1.28-1.28) | 1.16 (1.16-1.16) |
| 80-89 | 1.36 (1.36-1.36) | 1.13 (1.13-1.13) |
| 90+ | 1.18 (1.18-1.18) | 1.13 (1.13-1.13) |
| Sex |  |  |
| Male | Ref | Ref |
| Women | 1.65 (1.65-1.65) | 1.12 (1.12-1.12) |
| Education |  |  |
| 0-10 yrs | Ref | Ref |
| 11-15 yrs | 0.97 (0.97-0.97) | 0.97 (0.97-0.97) |
| 16+ yrs | 0.87 (0.87-0.87) | 0.98 (0.98-0.98) |
| Cohabitation status |  |  |
| Single | Ref | Ref |
| Married | 1.09 (1.09-1.09) | 0.95 (0.95-0.95) |
| Cohabitating | 1.07 (1.07-1.07) | 0.93 (0.93-0.93) |
| Immigration status |  |  |
| Danish born, Danish parents | Ref | Ref |
| Descendent of immigrant | 0.82 (0.82-0.82) | 0.87 (0.87-0.87) |
| Immigrant | 0.93 (0.93-0.93) | 0.88 (0.88-0.88) |
| Population density |  |  |
| 100-000+ | Ref | Ref |
| 20-000+ | 1.03 (1.03-1.03) | 1.00 (1.00-1.00) |
| 2000+ | 1.02 (1.02-1.02) | 1.01 (1.01-1.01) |
| <2,000 | 0.98 (0.98-0.98) | 1.01 (1.01-1.01) |
| Equivalised Income quintile |  |  |
| 1 | Ref | Ref |
| 2 | 1.08 (1.08-1.08) | 1.01 (1.01-1.01) |
| 3 | 1.08 (1.08-1.08) | 1.06 (1.06-1.06) |
| 4 | 1.07 (1.07-1.07) | 1.05 (1.05-1.05) |
| 5 | 1.08 (1.08-1.08) | 1.07 (1.07-1.07) |
| Intercept | 1.00 (1.00-1.01) | 67.29 (67.29-67.29) |

### eTable 9 Association between multimorbidity and OOP medicine expenditure (€) based on quantile regression

|  | Quantile 1 | Quantile 2 | Quantile 3 | Quantile 4 | Quantile 5 | Quantile 6 | Quantile 7 | Quantile 8 | Quantile 9 |
| --- | --- | --- | --- | --- | --- | --- | --- | --- | --- |
| No. of Conditions |  |  |  |  |  |  |  |  |  |
| 0 | Ref | Ref | Ref | Ref | Ref | Ref | Ref | Ref | Ref |
| 1 | 9.35 | 19.28 | 30.73 | 43.37 | 57.05 | 70.83 | 80.80 | 89.00 | 109.62 |
| 2 | 26.54 | 48.69 | 70.39 | 91.20 | 109.24 | 125.00 | 139.67 | 156.46 | 184.71 |
| 3 | 43.34 | 75.83 | 105.90 | 131.39 | 153.41 | 175.41 | 198.22 | 223.15 | 260.71 |
| 4 | 54.01 | 93.39 | 131.26 | 161.82 | 189.40 | 217.67 | 246.97 | 273.81 | 319.98 |
| 5+ | 67.32 | 112.12 | 161.06 | 203.21 | 243.71 | 282.98 | 318.88 | 357.81 | 407.88 |
| Age group |  |  |  |  |  |  |  |  |  |
| 18-29 | Ref | Ref | Ref | Ref | Ref | Ref | Ref | Ref | Ref |
| 30-39 | -1.82 | -2.42 | -1.74 | -0.51 | 0.90 | 3.70 | 8.56 | 15.01 | 23.20 |
| 40-49 | -1.57 | -1.64 | -0.06 | 2.41 | 5.96 | 10.99 | 17.84 | 21.94 | 28.59 |
| 50-59 | -0.18 | 1.20 | 4.55 | 8.73 | 13.60 | 19.34 | 25.33 | 25.83 | 31.76 |
| 60-69 | 1.45 | 3.95 | 7.85 | 12.68 | 17.34 | 22.89 | 28.50 | 28.04 | 31.35 |
| 70-79 | -3.06 | -4.16 | -2.08 | 1.59 | 5.98 | 11.19 | 16.74 | 17.29 | 19.32 |
| 80-89 | -6.43 | -9.09 | -6.99 | -2.89 | 1.66 | 7.71 | 12.31 | 11.84 | 11.99 |
| 90+ | -6.35 | -6.98 | -3.74 | 1.62 | 6.39 | 12.46 | 16.36 | 14.33 | 13.62 |
| Sex |  |  |  |  |  |  |  |  |  |
| Male | Ref | Ref | Ref | Ref | Ref | Ref | Ref | Ref | Ref |
| Women | 2.62 | 5.21 | 7.37 | 9.92 | 12.81 | 15.40 | 17.25 | 16.66 | 5.81 |
| Education |  |  |  |  |  |  |  |  |  |
| 0-10 yrs | Ref | Ref | Ref | Ref | Ref | Ref | Ref | Ref | Ref |
| 11-15 yrs | 1.00 | 1.63 | 1.63 | 1.08 | 0.27 | -1.05 | -3.24 | -6.42 | -12.59 |
| 16+ yrs | 0.76 | 1.36 | 1.58 | 1.33 | 1.16 | 0.66 | -0.42 | -3.01 | -9.94 |
| Cohabitation status |  |  |  |  |  |  |  |  |  |
| Single | Ref | Ref | Ref | Ref | Ref | Ref | Ref | Ref | Ref |
| Married | 0.35 | 0.50 | 0.34 | -0.48 | -1.73 | -3.31 | -6.20 | -9.61 | -19.37 |
| Cohabitating | -0.51 | -1.22 | -2.08 | -3.54 | -5.56 | -7.45 | -10.90 | -14.67 | -21.23 |
| Immigration status |  |  |  |  |  |  |  |  |  |
| Danish born, Danish parents | Ref | Ref | Ref | Ref | Ref | Ref | Ref | Ref | Ref |
| Descendent | -1.48 | -2.85 | -4.35 | -6.57 | -9.79 | -14.12 | -20.40 | -27.99 | -31.67 |
| Immigrant | -0.68 | -0.95 | -1.08 | -2.03 | -3.82 | -7.04 | -12.92 | -23.76 | -27.36 |
| Population Density |  |  |  |  |  |  |  |  |  |
| 100,000+ | Ref | Ref | Ref | Ref | Ref | Ref | Ref | Ref | Ref |
| 20,000+ | -0.25 | -0.16 | -0.03 | 0.32 | 0.62 | 0.97 | 1.28 | 0.82 | 0.86 |
| 2,000+ | -0.14 | -0.03 | 0.09 | 0.41 | 0.67 | 0.82 | 1.14 | 0.83 | 0.63 |
| <2,000 | -0.34 | -0.40 | -0.42 | -0.29 | -0.21 | 0.00 | 0.37 | 0.43 | 0.17 |
| Income quintile |  |  |  |  |  |  |  |  |  |
| 1 | Ref | Ref | Ref | Ref | Ref | Ref | Ref | Ref | Ref |
| 2 | 1.07 | 1.25 | 1.19 | 1.32 | 1.56 | 1.47 | 1.84 | 2.34 | 2.83 |
| 3 | 3.90 | 5.93 | 7.42 | 9.25 | 10.15 | 10.67 | 10.86 | 10.50 | 7.91 |
| 4 | 3.25 | 4.68 | 5.47 | 6.58 | 6.85 | 6.63 | 6.21 | 5.82 | 2.43 |
| 5 | 2.93 | 4.06 | 4.68 | 5.77 | 5.90 | 5.93 | 5.78 | 5.34 | 3.58 |
| Intercept | 6.03 | 9.06 | 12.97 | 18.85 | 28.60 | 43.55 | 68.87 | 116.37 | 198.16 |

^** p<.05; * p<.001^
